# An effect of the COVID-19 pandemic: significantly more complicated appendicitis due to delayed presentation of patients!

**DOI:** 10.1101/2021.01.23.21250358

**Authors:** Marie Burgard, Floryn Cherbanyk, Konstantinos Nassiopoulos, Sonaz Malekzadeh, François Pugin, Bernhard Egger

## Abstract

**AIMS OF THE STUDY:** The novel coronavirus pandemic has affected emergency department consultations for surgical pathologies. The aim of our study was to compare the number of acute appendicitis cases and the proportion of complicated appendicitis before and during the COVID-19 pandemic.

**METHODS:** We retrospectively analyzed all data collected from a multi-center database of patients presenting to the emergency department for acute appendicitis during the COVID-19 pandemic from March 12 to June 6, 2020, and compared these data with those from the same periods in 2017, 2018, and 2019. The number of acute appendicitis cases, proportion of complicated appendicitis, and pre- and postoperative patient characteristics were evaluated.

**RESULTS:** A total of 306 patients were included in this evaluation. Sixty-five patients presented during the 2020 COVID-19 pandemic lockdown (group A), and 241 patients in previous years (group B: 2017–2019). The number of consultations for acute appendicitis decreased by almost 20 percent during the pandemic compared with previous periods, with a significant increase in complicated appendicitis (52% in group A versus 20% in group B, p < 0,001.). Comparing the two groups, significant differences were also noted in the duration of symptoms (symptoms > 48h in 61% and 26%, p < 0,001), the intervention time (77 vs 61 minutes, p = 0,002), length of hospital stay (hospitalization of > 2 days in 63% and 32%, p < 0.001) and duration of antibiotic treatment (antibiotics > 3 days in 36% and 24% p = 0.001).

**CONCLUSIONS:** The COVID-19 pandemic resulted in a decreased number of consultations for acute appendicitis, with a higher proportion of complicated appendicitis, most likely due to patient delay in consulting the emergency department at symptom onset. Patients and general practitioners should be aware of this problem to avoid a time delay from initial symptoms to consultation.

## Introduction

The novel coronavirus SARS-CoV-2 (causing COVID-19) was first recognized in China in December 2019 and rapidly spread throughout the world. The COVID-19 disease outbreak was declared a pandemic by the World Health Organization (WHO) on March 11, 2020 [1]. Because of the high contagious potential of this virus, many hospitals quickly experienced patient overload and an increased need to use respiratory assistance in infected patients. Consequently, many countries announced a state of emergency and advised the population to stay at home whenever possible. However, patients’ fear of becoming infected by the virus obviously also led to a marked reduction in consultations at emergency departments, even in the case of abdominal symptoms [2].

Appendicitis is one of the most common causes of acute abdominal pain and is one of the most frequent reasons for emergency surgery [3]. Complicated appendicitis is defined as gangrenous or perforated appendicitis or peri-appendicular abscess formation. Longer duration of symptoms prior to consultation is a well-known risk factor for developing complicated appendicitis [4].

During the COVID-19 pandemic, the number of consultations for acute appendicitis decreased. While some studies demonstrated a higher rate of complicated appendicitis, probably due to delayed consultation, other studies have reported no change in the rates of uncomplicated/complicated appendicitis during this period [5–8].

The aim of our study was to analyze the incidence of acute appendicitis as well the rates of uncomplicated/complicated appendicitis during the COVID-19 lockdown period and to compare these data with a comparable period in previous years. We found that the COVID-19 pandemic resulted in a decreased number of consultations for acute appendicitis, a longer delay to first consultation, a higher proportion of complicated appendicitis, longer use of post-surgery antibiotic treatment, and longer hospital stay, most likely due to patient delay in consulting the emergency department at symptom onset.

## Materials and methods

Data from patients was extracted from prospective digital databases of a multi-site tertiary care hospital (HFR, Cantonal Hospital of Fribourg) and a secondary care hospital (Daler Hospital). The retrospective analysis included all patients who presented at the respective EDs for acute appendicitis during the COVID-19 lockdown period in Switzerland from March 12 to June 6, 2020 (group A), and for comparison data of patients presenting for the same reason and during the same period of the year in 2017, 2018, and 2019 (group B).

### Data analysis

Demographic data, biologic and inflammatory markers at presentation, duration of symptoms until presentation to the ED, duration of antibiotic treatment, and duration of hospital stay were recorded. Each operation protocol was separately evaluated and the severity of appendicitis classified according to the laparoscopic grading score of Gomes [9]: briefly, normal-looking appendix = grade 0, non-complicated appendicitis with redness and edema = grade 1, with additional fibrin layers = grade 2, complicated appendicitis with segmental peripheral necrosis = grade 3A, with necrosis of the appendix base = grade 3B, with abscess formation = grade 4A, with regional peritonitis = grade 4B, and with diffuse peritonitis = grade 5.

In addition, histopathologic results were recorded and classified into five groups: grade 0 = no inflammation, grade 1 = acute appendicitis, grade 2 = ulcero-phlegmonous appendicitis, grade 3 = necrotizing or gangrenous appendicitis, grade 4 = abscess formation, and grade 5 = perforated appendicitis. Grades 1 and 2 were classified as non-complicated and grades 3–5 as complicated appendicitis.

Finally, patient data were compared between group A (COVID lockdown, March 12–June 6, 2020) and group B (March 12–June 6, 2017, 2018, and 2019).

### Statistical analysis

Statistical analysis was performed utilizing the Statistical Package for Social Sciences, version 27.0 (SPSS, IBM, Armonk, USA). Quantitative variables were expressed as mean ± standard deviation (SD) and ranges. Qualitative variables were expressed as raw numbers, proportions, and percentages. Pearson’s chi-square test was used to search for differences between categorical variables while the Student t-test and Mann-Whitney U test were used for continuous variables. All results were expressed with a 95% confidence interval, and statistical significance was defined as a *P* value <0.05.

This study was approved by the local Ethics Committee (Project-ID 2020-01676).

## Results

### Number of appendicitis cases

A total of 311 patients presented to the respective EDs for suspicion of acute appendicitis during the reviewed periods. Five patients with a different peri-operative diagnosis were excluded from analysis. In all, 306 patients were enrolled, including 65 who presented during the COVID-19 lockdown and 245 in the years before (80 patients in 2017, 78 in 2018, and 87 in 2019). On an annual basis, the total number of patients presenting with acute appendicitis during the COVID-19 pandemic decreased by almost 20% when compared with the years before.

### Patients’ pre-operative characteristics

There were no statistically significant differences in median age and sex distribution between the groups. In 12 patients, the duration of symptoms prior to consultation could not be re-evaluated. However, most of the patients in group A (61%) presented at the ED more than 48 h after the beginning of symptoms, compared with 26% in group B (*P* < 0.001). Regarding the biologic inflammatory markers at presentation, plasma C-reactive protein (CRP) was significantly higher in group A compared with group B (*P* < 0.007), while the white blood cell count (WBC) was only slightly and nonsignificantly higher in group A (*P* = 0.438) when compared with group B. The demographic, clinical, and biological data of the two groups are summarized in Table 1.

**Table 1:**
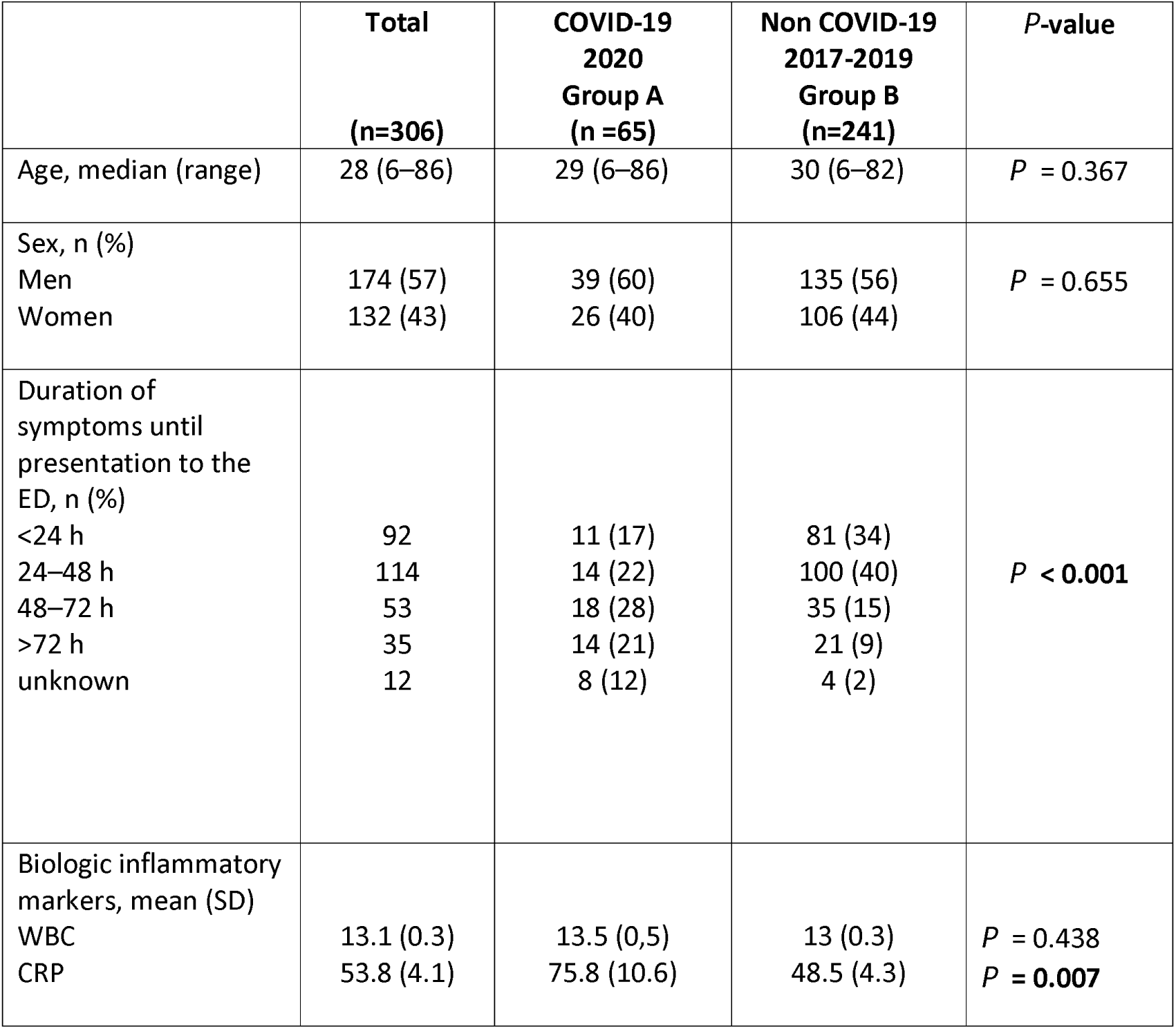
Patients’ demographic and preoperative characteristics n= number; ED = emergency department; h = hour s; SD = standard deviation; WBC = white blood cell count; CRP = C-reactive protein

### Operative and postoperative characteristics

Six patients (2%) were treated conservatively, one patient during the COVID-19 period and five patients in group B. Among them, five patients presented with complicated appendicitis diagnosed on imaging and were treated using antibiotic therapy and transabdominal drainage. One patient with a non-complicated appendicitis and multiple comorbidities underwent antibiotic treatment only.

One patient was operated on using an open McBurney approach; the remainder all underwent a laparoscopic intervention (Table 2).

**Table 2:**
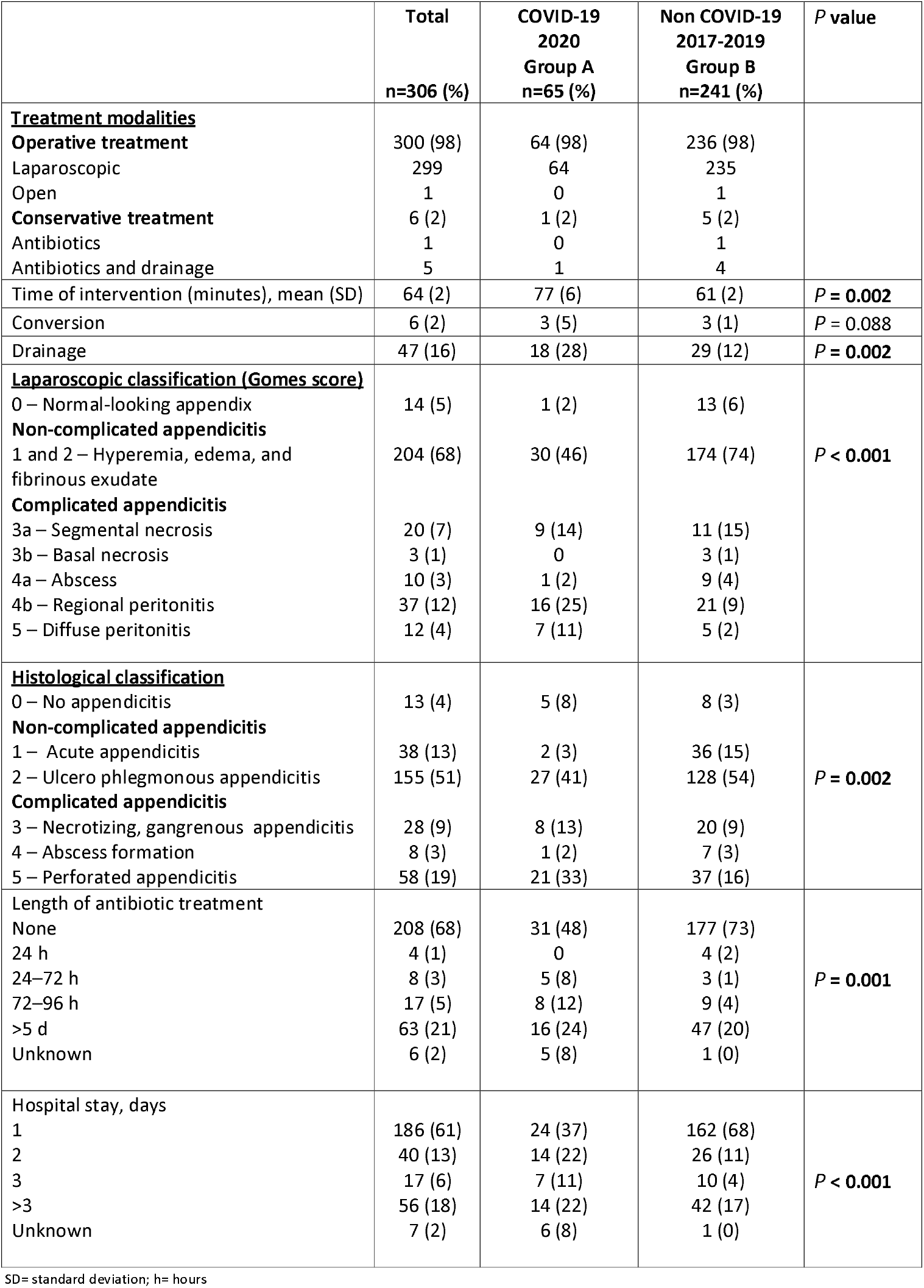
Operative and postoperative characteristics

A significantly higher proportion of complicated appendicitis was observed in group A, based on the Gomes and histopathological classifications being 52% and 48% in group A, and 20% and 28% in group B. The rates of non-complicated and complicated appendicitis over the last 4 years in both groups are illustrated in Fig 1.

**Fig 1.**
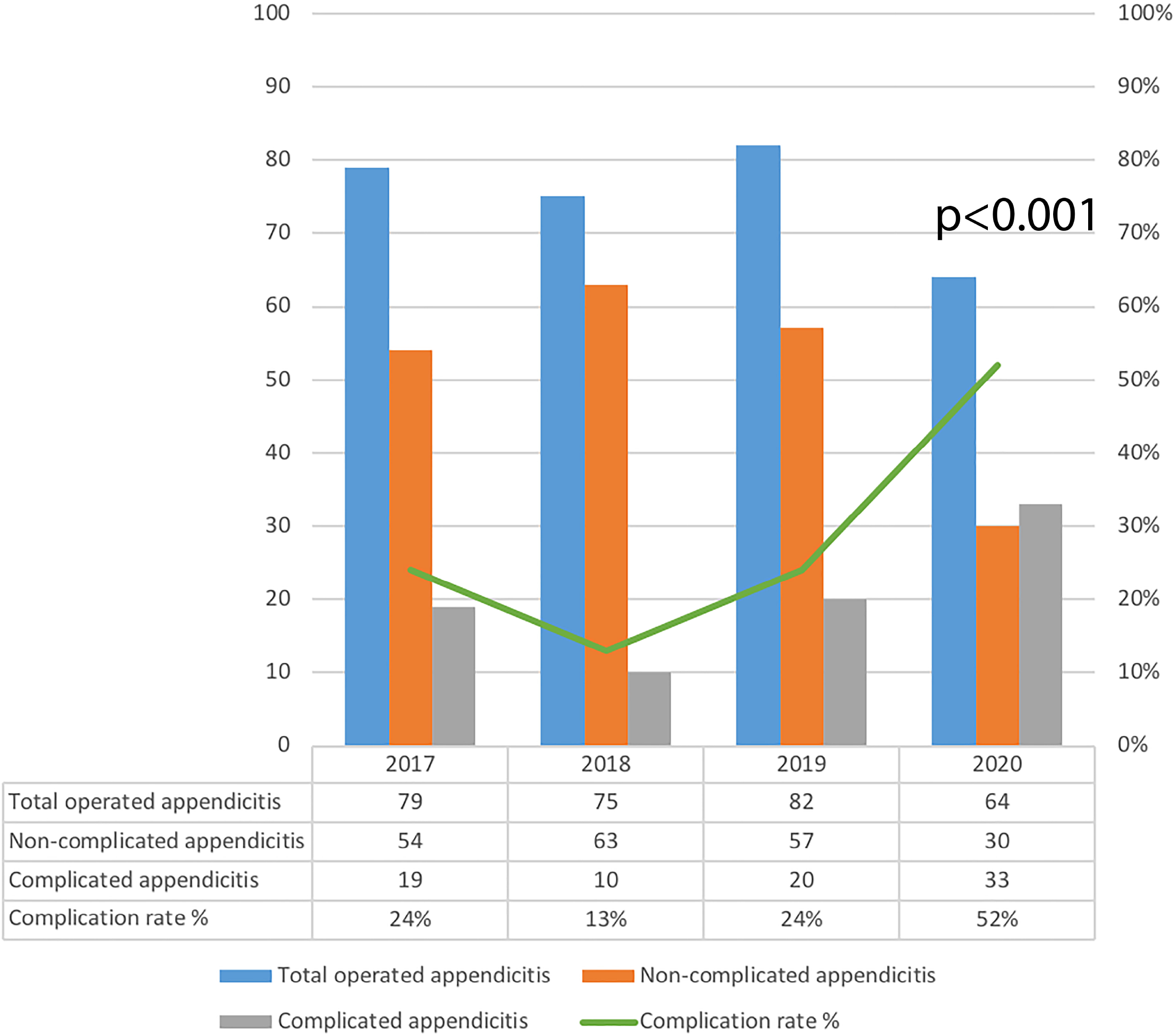
Distribution of complicated and non-complicated appendicitis. Distribution of non-complicated and complicated appendicitis based on the laparoscopic grading score. Normal-looking appendix is not included in the graphic.

Patients in the COVID-19 group had a significantly longer intervention time, 77 vs. 61 minutes (mean, *P* = 0.002), and a higher rate of transabdominal drainage, 28% vs. 12%, (*P* = 0.002) in group A vs. B, respectively. Post surgery, there was a longer need for antibiotics in group A compared with group B patients (*P* = 0.001). The percentages of patients who underwent antibiotic therapy for more than 3 days were 36% and 24% in group A and B, respectively (*P* = 0.001). Information regarding the duration of antibiotic treatment was not available for five patients in group A.

In addition, patients in group A experienced a significantly longer hospital stay when compared with patients of group B: 63% and 32% of patients were hospitalized ≥2 days in groups A and B, respectively (*P* < 0.001).

## Discussion

The present study demonstrates a diminished rate of patients presenting at the ER with acute appendicitis, a longer delay to first consultation, a higher rate of complicated appendicitis, and a prolonged hospital stay in patients presenting during the COVID-19 pandemic period when compared with previous years.

The incidence of acute appendicitis during the COVID-19 pandemic is comparable to the findings of other reported studies [5–7, 10–12]. The encouragement by authorities to stay at home whenever possible, along with fear of being contaminated by the virus, might explain the fact of fewer consultations at emergency departments. Additionally, some spontaneous resolution of the disease might further explain the reduced rate of patients with diagnosis of acute appendicitis during the COVID-19 period. Seasonal changes and a tendency to a decreasing incidence of acute appendicitis over the years have also also reported as possible factors responsible for this finding [11, 13]. However, because in the present study the same seasonal periods over consecutive years were compared, it is unlikely that seasonal changes affected the incidence of acute appendicitis. A trend of fewer patients with acute appendicitis during the 3 previous years has not been confirmed by our evaluation.

Pre-operative investigations of the studied patients demonstrated a significantly higher CRP value at presentation in affected patients during the COVID-19 period when compared with previous time periods. Our study also demonstrated that the majority of patients (61%) admitted during the COVID-19 period presented later than 48 hours after the beginning of symptoms, compared with only 26% in previous years. A lower consultation rate and a higher delay to first consultation has been reported to be directly related to a higher rate of complicated appendicitis in such patients [14]. Our study confirms this finding, with significantly more patients with complicated appendicitis during the COVID-19 period when compared with previous years. This is in accord with other published reports [6–8, 10, 15]. However, Tankel et al. [5] did not observe different rates of non-complicated and complicated appendicitis during the COVID-19 pandemic. They therefore concluded that some patients with beginning appendicitis might possibly be treated conservatively.

Our results show a longer mean intervention time during the COVID-19 lockdown that is easily explained by the higher rate of complicated appendicitis. In addition, most of our patients admitted during the lockdown period had a significantly longer hospital stay when compared with patients of the pre-pandemic group. Furthermore, prolonged post-operative antibiotic treatment during the COVID-19 period was noted with respect to the previous years, and this is also explained by the higher rate of complicated appendicitis. Interestingly, these results do not confirm those of other studies [6, 7], where operation time and the post-operative recovery period were not different during the COVID-19-period (first wave) when compared with previous years.

Our study has some limitations. First, the period from March 12 to June 6 was chosen according to the coronavirus lockdown period in Switzerland. Choosing an earlier end date of the study might have led to different results. Second, general practitioners might have treated some patients conservatively without referring them to an emergency department, resulting in a possible underestimation of the total number of patients with appendicitis during this period. However, in our area general practitioners usually send most patients with a suspicion of acute appendicitis to the emergency department in order that they undergo imaging studies to confirm the suspected diagnosis. Thus, we believe that the number of patients who did not present to one of the participating hospitals was rather low.

In conclusion, the COVID-19 pandemic induced a decrease in acute appendicitis cases in the EDs, with a significant increase in complicated appendicitis. This phenomenon is most likely explained by symptomatic patients’ substantially delaying medical consultation. The observed increased number of patients with complicated appendicitis was associated with a longer intervention time, longer antibiotic treatment, and a longer hospital stay compared with pre-pandemic patients. Since there might be another lockdown due to COVID-19 or other pandemics in future, it would be helpful to inform patients and general practitioners broadly of the importance of avoiding time delay from onset of symptoms to consultation.

## Data Availability

Data not available due to [ethical/legal/commercial] restrictions

## Acknowledgments

We thank the surgical staff of the Daler hospital for the scientific collaboration.

The authors declare that they have no competing interests.

